# Effects of sodium intake reduction on blood pressure control in hypertensive patients

**DOI:** 10.1101/2023.09.18.23295627

**Authors:** Ronak Brijeshkumar Upadhyay, Binay K Panjiyar, Abhimanyu Agarwal, Dhwani Manishbhai Patel, Monica Ghotra, Gulam Husain Nabi Husain Mahato

## Abstract

The primary objective of this research was to delve into the impact of sodium reduction on managing blood pressure in individuals with hypertension. The investigation entailed a thorough examination of pertinent literature, concentrating on experimental inquiries, meta-analyses, and clinical studies spanning the years 2013 to 2022. The focus encompassed an array of scholarly works that go through the intricate link between sodium consumption and the regulation of blood pressure. These diverse studies yielded noteworthy outcomes, consistently portraying a proportional response to salt reduction. Notably, the reduction in sodium intake showcased a significant reduction in blood pressure levels among hypertensive patients. The underpinnings of this phenomenon were scrutinized, encompassing variations in renin, aldosterone, catecholamine, cholesterol, and triglyceride levels. Furthermore, these studies give the interplay of salt reduction with dietary interventions and the baseline blood pressure, accentuating the potential for custom-tailored methodologies to optimize blood pressure management. In recent times, lifestyle interventions targeting sodium intake have gained escalated consideration, and the effectiveness of these interventions in lowering blood pressure among hypertensive patients has been substantiated through randomized controlled trials. These findings underscore the pivotal role of curtailing sodium intake as a potent strategy for controlling blood pressure effectively in the hypertensive demographic. Nonetheless, further exploration is warranted to delve into prolonged outcomes, refine dietary recommendations, and formulate comprehensive strategies for fine-tuning blood pressure control through judicious sodium reduction.

## Introduction and Background

Hypertension, a prevalent cardiovascular ailment characterized by consistently high blood pressure levels, continues to pose a significant global public health challenge. It stands as a primary risk factor for a range of unfavorable consequences, including stroke, heart attacks, cardiac insufficiency, and renal impairment. The World Health Organization (WHO) has estimated that hypertension significantly contributes to the global disease burden and underscores its considerable public health implications [4].

Within the realm of hypertension’s development and management, dietary factors play a pivotal role. Among these, excessive sodium intake has emerged as a pivotal modifiable element. Sodium, a fundamental electrolyte, serves essential roles in numerous physiological processes such as maintaining fluid equilibrium, nerve function, and muscle contraction. However, when sodium intake surpasses optimal levels—often through table salt consumption—it invariably corresponds with heightened blood pressure and escalated cardiovascular risks [1][4].

Extensive research has dissected the intricate interaction between sodium intake and the intricate regulation of blood pressure, underscoring the significance of interventions aimed at reducing salt intake in hypertension management. Consistently, investigations highlight a dose-dependent correlation between sodium consumption and blood pressure. Particularly noteworthy is a comprehensive dose-response meta-analysis by Filippini et al. (2021) that condenses an experimental exploration showcasing the extent of blood pressure modulation due to sodium depletion [1]. Furthermore, Graudal et al. (2020) meticulously conducted a systematic review and meta-analysis investigating the ramifications of high and low sodium diets on blood pressure as well as factors like renin, aldosterone, catecholamines, cholesterol, and triglycerides [2]. The synthesis of these inquiries amplifies the potential efficacy of salt reduction as a pivotal avenue for orchestrating blood pressure control.

As the reservoir of evidence favoring salt reduction burgeons, there is a growing inclination toward personalized strategies in managing hypertension. The success of salt-reduction interventions might be influenced by variables including initial blood pressure readings, genetic predisposition, and the individual’s responses to dietary alterations. In a notable instance, Juraschek et al.’s (2017) investigation scrutinized the effects of curtailed sodium intake and the Dietary Approaches to Stop Hypertension (DASH) diet—designed to combat hypertension—in relation to baseline blood pressure, underscoring the pertinence of personalized attributes in crafting treatment plans [6][7].

While the broad benefits of salt reduction remain evident, there is a need for in-depth exploration, especially concerning hypertensive individuals. This led Shamsi et al. (2021) to embark on a randomized controlled trial that delved into the repercussions of lifestyle interventions on curbing dietary sodium intake and modulating blood pressure among hypertensive patients, yielding pragmatic insights into the real-world application of salt reduction strategies [8].

Within this context, the present systematic review strives to comprehensively assess the implications of salt reduction in blood pressure management among hypertensive patients. By amalgamating evidence from a spectrum of sources including experimental studies, meta-analyses, and randomized controlled trials, this review endeavors to offer a nuanced comprehension of the interplay between salt reduction and blood pressure outcomes in the realm of hypertension. The ensuing section undertakes an exhaustive scrutiny of the chosen studies, focusing on methodologies, interventions explored, and their potential ramifications for clinical practice and public health paradigms.

## Review

### Methods

The aim for this research was to focus on the recent studies concerning specifically to control blood pressure in hypertensive patients with limiting or restricting the salt intake. We didn’t include the papers which were older than 2013 for getting the latest advancements achieved in this topic. This systematic review follows the latest guidelines of preferred Reporting Items for Systematic Reviews and Meta-analysis (PRISMA) issued in 2020. The data collected from published papers and eliminating the need for ethical approval is shown in figure 1.

**Figure 1:**
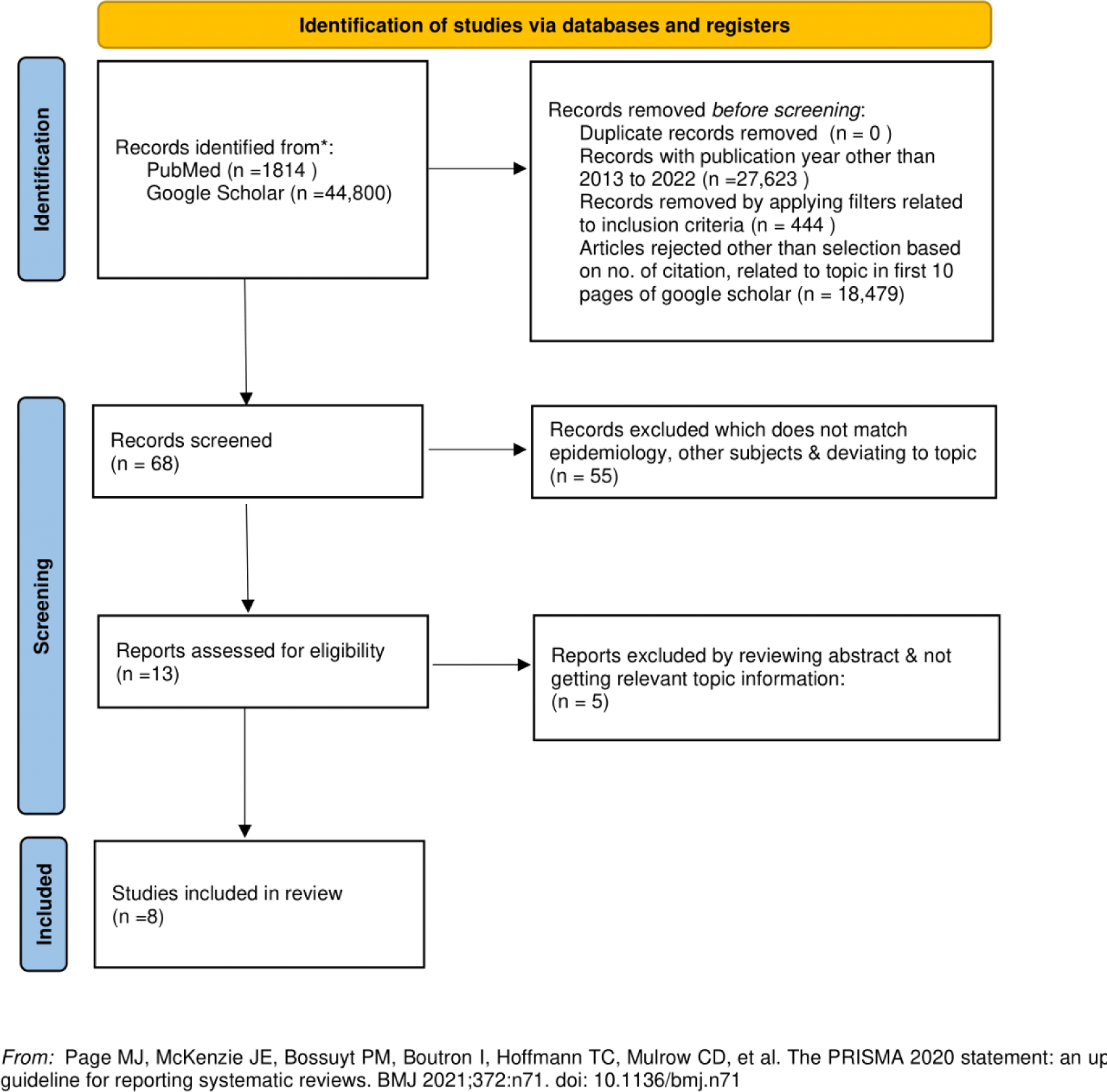
PRISMA flow diagram illustrating the search strategy and study selection process for the systematic review.

### Literature search and study selection

For the systematic review, we decided and searched for relevant papers according to keywords by using databases PubMed including Medline and google scholar for broad search. We used all the databases simultaneously and searched for the studies that were mentioned in review papers, editorials, and comments on the search databases. We continued our search for additional studies that satisfied our inclusion criteria.

After searching in databases, gathered a list of abstracts that were then reviewed independently for further selection using our inclusion criteria. Moreover, our aim was to gather the studies that majorly involved human studies, English language, age of 19 and more, and excluding the studies which doesn’t have relevant information to our title.

### Inclusions & Exclusions criteria

To start with the study, we defined some inclusion and exclusion criteria and followed them throughout our search strategy and study selection. Majorly the study selection was through inclusion criteria but exclusion criteria helped out to filter the studies which weren’t relevant for our study.

### Search strategy

The search was conducted on PubMed including Medline and google scholar library using the relevant keywords for our title. Boolean operators such as “AND” & “OR” were used in the databases. AND for covering all the keywords which aren’t similar/synonym to each other and sustain the essential ideology of the title. OR was used for the similar keywords who are related to each other and have almost similar meaning. Using the Boolean operators, we searched and gathered the initial number of studies related to our title. A detailed and comprehensive search strategy is mentioned below in Table 2.

**Table 1:** Inclusions and Exclusions criteria utilized during the literature review search.

| Inclusion | Exclusion |
| --- | --- |
| The study should mainly involve the Humans, whether male or female. | Studies on Animals |
| Studies conducted on patients with history of Hypertension | Studies comprising of patients with other disease |
| The timeline should be of last 10 years i.e., (01/01/2013 to 12/31/2022) | Studies published before 2013 |
| Studies which can be accessed as Free full text | Paid papers |
| Studies which were strictly in English language | Pregnant women |

**Table 2:**
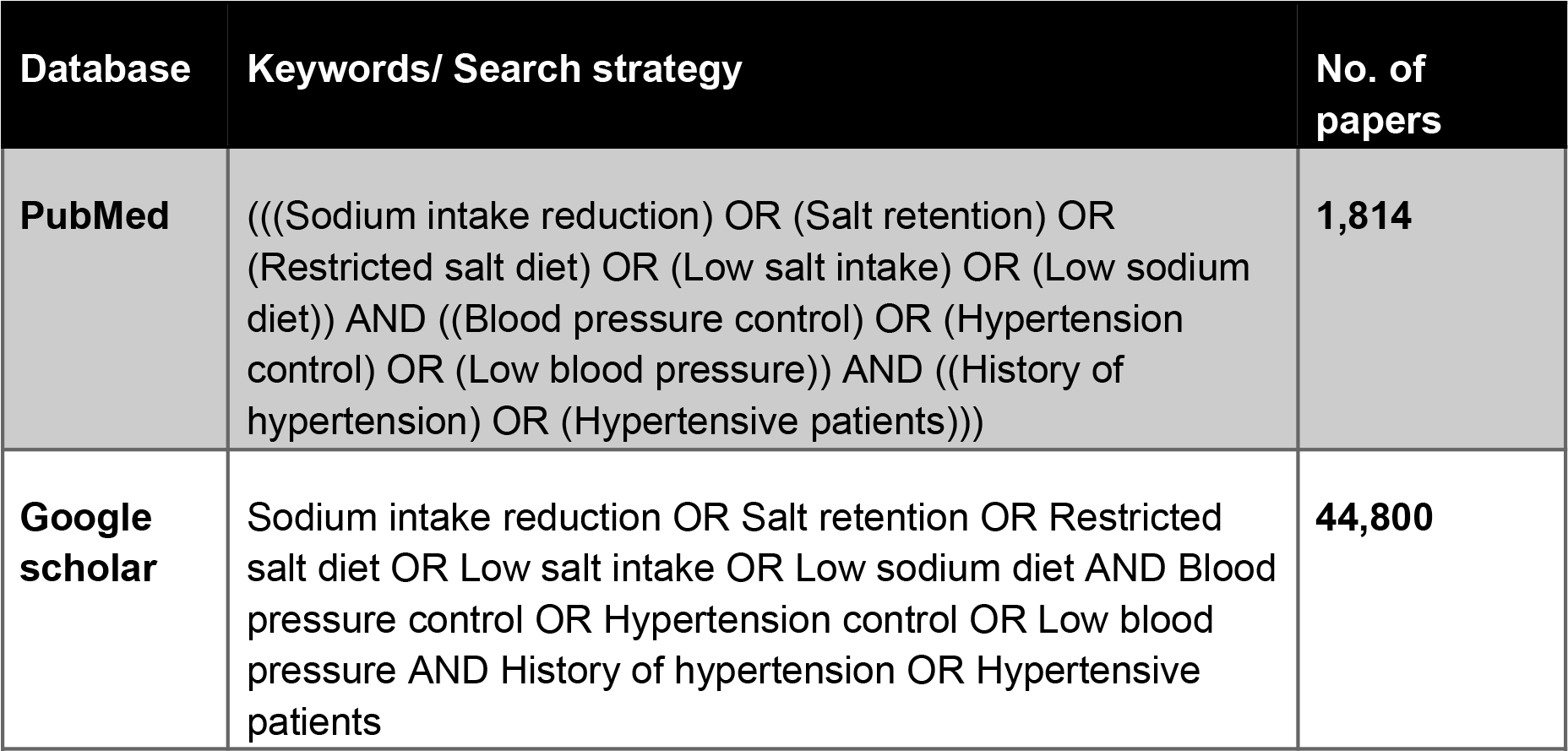
Search strategy with keywords to acquire number of papers for literature search from databases.

### Quality appraisal

We used various quality assessment tools to ensure the authenticity of the selected works. For systematic review and meta-analysis, we used the PRISMA checklist and Cochrane bias tool assessments for randomized clinical trials. Nonrandomized clinical trials were evaluated using the Newcastle-Ottawa Instrument Scale. We assessed the quality of the qualitative studies using the Critical Appraisal Skills Program (CASP) checklist, as shown in Table 3. To avoid confusion in taxonomy, we used the Narrative Review Article Rating Scale (SANRA) to rate article quality.

**Table 3:** Showing quality appraisal tools used. **PRISMA: Preferred reporting items for systematic reviews and meta-analyses; SANRA: Scale for the assessment of non-systematic review articles**

| Quality Appraisal Tools Used | Type of Studies |
| --- | --- |
| Cochrane Bias Tool Assessment | Randomized Control Trials |
| Newcastle-Ottawa Tool | Non-RCT and Observational Studies |
| PRISMA Checklist | Systematic Reviews |
| SANRA Checklist | Any Other Without Clear Method Section |

## Results

After conducting comprehensive searches in two chosen databases, namely PubMed and Google Scholar, we identified a total of 46,614 articles. Our subsequent meticulous review process involved the application of specific criteria, resulting in the exclusion of 46,546 articles. Among the initial pool, 68 papers remained for detailed assessment; however, 55 of these were not considered suitable due to their focus on epidemiology, different subject matter, or straying from the central topic. A thorough examination of the remaining 13 papers revealed that five of them exhibited inadequate titles and abstracts, leading to their exclusion. Finally, a rigorous quality assessment was performed on the remaining eight papers, all of which aligned with our predetermined criteria. This final set of eight articles constitutes the corpus of our systematic review. Further insights into each of these articles are elaborated in Table 4.

**Table 4:**
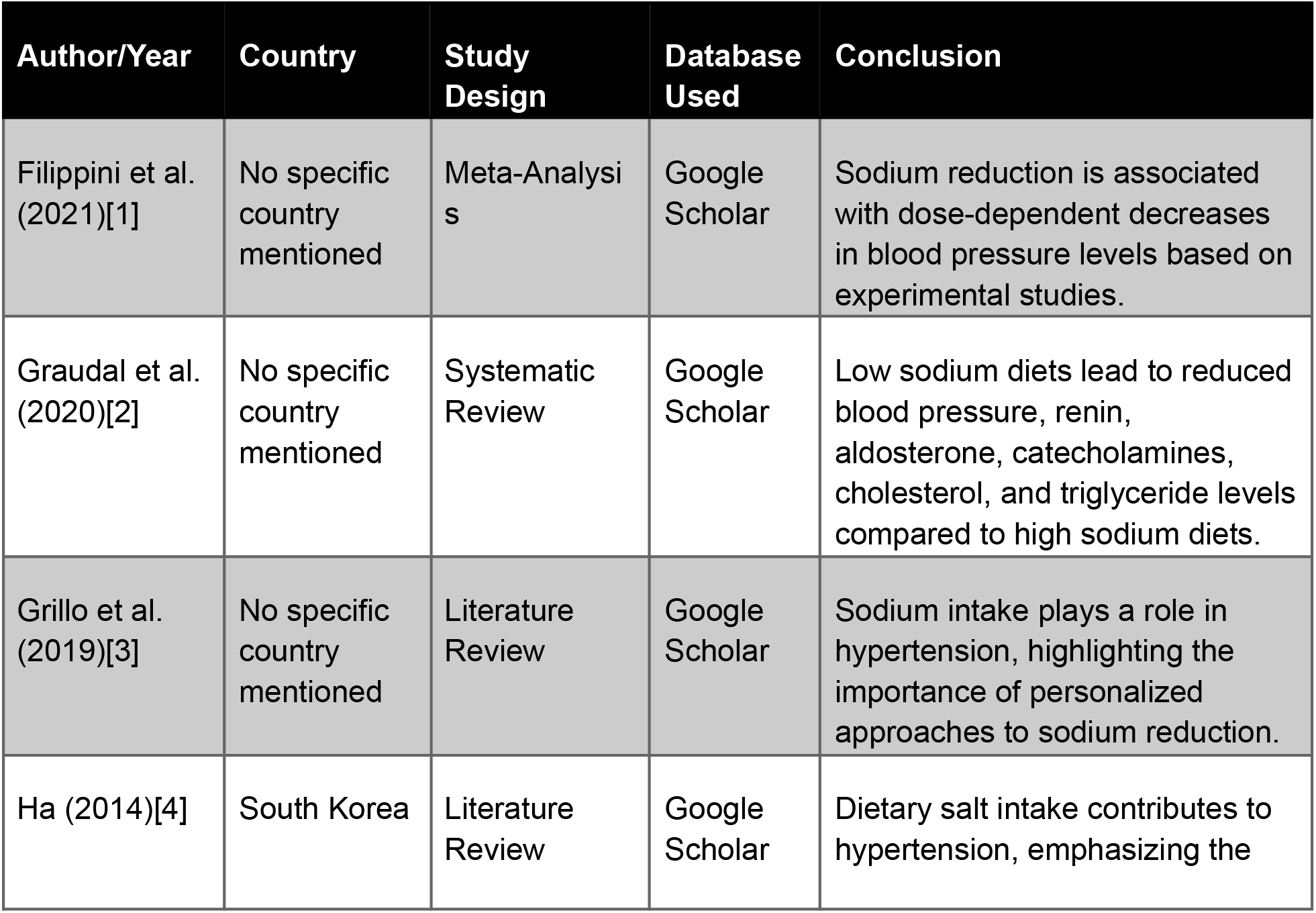
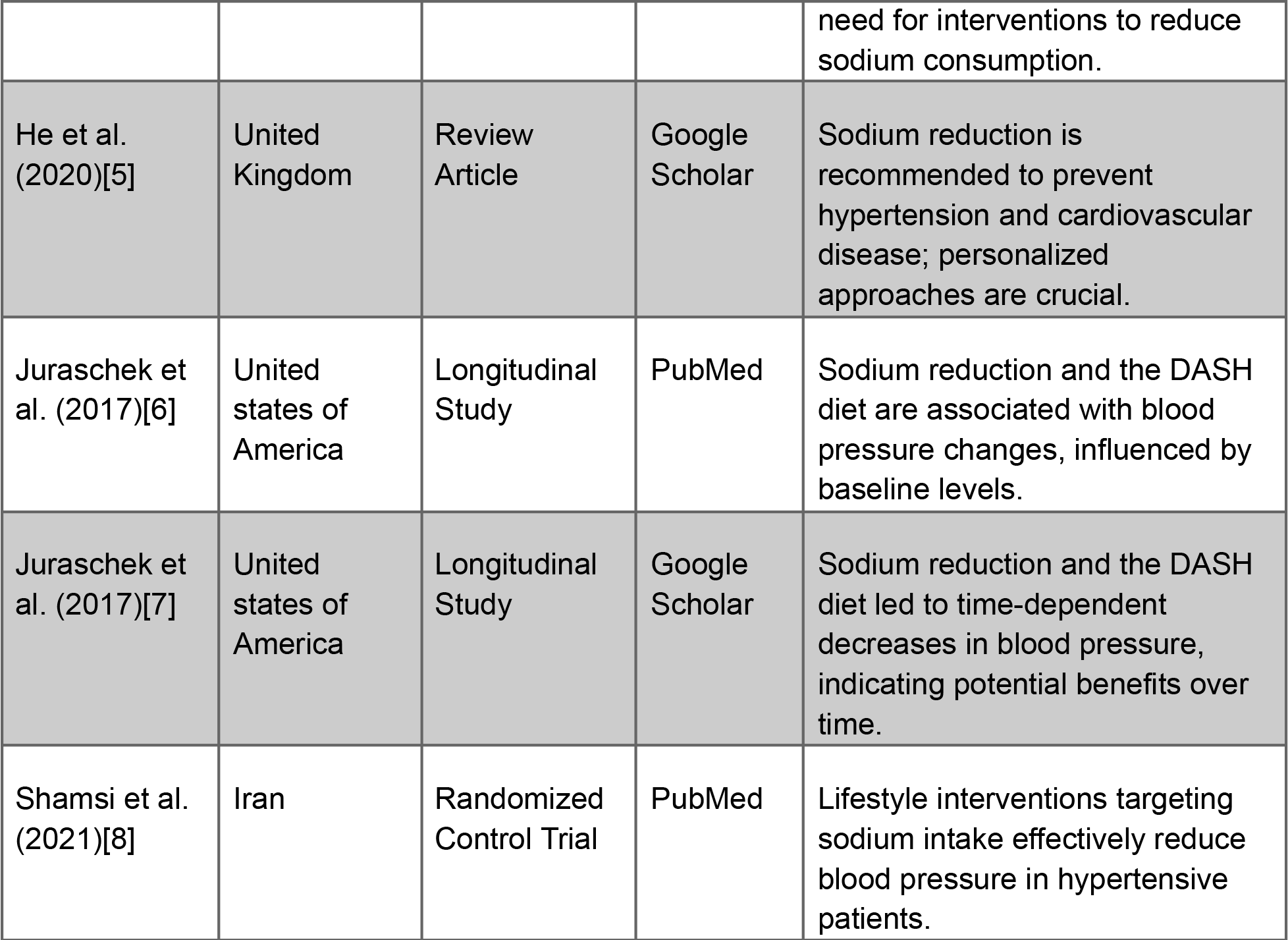
Summary of the results of the selected papers.

## Discussion

A comprehensive exploration of the impact of reducing sodium intake on the management of blood pressure among individuals with hypertension, incorporating insights from a diverse array of studies, provides valuable perspectives on the potential benefits and complexities associated with salt reduction strategies. The endeavor to lower sodium intake as a means to control blood pressure in hypertensive individuals holds substantial significance, underpinned by a substantial body of evidence from various research endeavors. These collective findings offer valuable insights into the intricate interplay between sodium reduction and its effects on blood pressure regulation.

Foremost, multiple investigations consistently affirm that the reduction of sodium intake leads to a decrease in blood pressure, underscoring the significance of this dietary adjustment [1][2][3]. While the extent of blood pressure reduction varies between systolic and diastolic measurements, one study reveals that the reduction in systolic blood pressure (SBP) is notably more pronounced than that in diastolic blood pressure (DBP) when sodium intake is curtailed by less than 2 grams per day. This divergence becomes more conspicuous when sodium reduction is achieved through initial sodium restriction followed by behavioral interventions. Intriguingly, there seems to exist a threshold effect for DBP, albeit not for SBP, when sodium intake is reduced by less than 2 grams per day, shedding light on the nuanced connection between sodium reduction and blood pressure control [1].

Salt sensitivity, characterized by varying blood pressure responses to alterations in salt intake, emerges as a pivotal factor in hypertension [3]. Hypertensive individuals tend to experience more pronounced reductions in blood pressure when sodium intake is curtailed, further underscoring the relevance of sodium reduction for those with hypertension [2]. Excessive salt consumption has been unequivocally linked to the onset of hypertension and the concomitant cardiovascular complications, cementing the imperative nature of controlling sodium intake [3].

On a global scale, sodium intake has witnessed a dramatic surge, with contemporary average consumption levels far surpassing historical norms [4][5]. This upsurge in sodium intake carries substantial health ramifications, contributing significantly to the burden of disability-adjusted life-years and mortality, particularly attributed to cerebrovascular and ischemic heart diseases [4][5]. Recognizing this, esteemed health organizations such as the World Health Organization (WHO) advocate for population-wide salt reduction as an indispensable public health measure [4].

The Dietary Approaches to Stop Hypertension (DASH) diet, characterized by its emphasis on fruits, vegetables, and low-fat dairy products while concurrently reducing saturated fat and cholesterol, has been empirically validated as an effective approach for reducing both SBP and DBP [6]. The amalgamation of the DASH diet with sodium reduction amplifies its beneficial effects, resulting in even more pronounced reductions in blood pressure, especially among individuals with elevated baseline SBP [6].

It’s important to acknowledge that the full potential of long-term sodium reduction may not be immediately realized, underscoring the significance of sustained efforts to maintain lower blood pressure levels [7]. Additionally, combining sodium reduction with diets rich in potassium can augment its advantages, emphasizing the interconnected nature of dietary choices in influencing blood pressure [7].

Lifestyle interventions, particularly Comprehensive Care Models (CCM), have demonstrated their effectiveness in enhancing patient adherence to dietary sodium restrictions among hypertensive individuals [8]. These interventions play a pivotal role in bolstering sodium intake reduction and, consequently, reducing both SBP and DBP [8].

Furthermore, an examination of the dose-response relationship between salt intake and blood pressure, based on two well-controlled trials, provides compelling evidence that lower salt intake is associated with lower blood pressure levels [5]. This aligns with epidemiological studies and experiments in chimpanzees, suggesting that further reducing salt intake to levels even lower than those recommended by the World Health Organization may yield greater health benefits [5].

The collective body of evidence paints a coherent picture of the significance of sodium reduction as a fundamental strategy for blood pressure control, particularly among individuals with hypertension. It should serve as a cornerstone of public health initiatives aimed at mitigating the risks associated with elevated blood pressure and related cardiovascular diseases. These findings underscore the intricate relationship between dietary choices, lifestyle interventions, and adherence to recommended sodium intake levels in promoting improved cardiovascular health, especially in the context of the global surge in sodium consumption.

### Limitations

The literature review is limited due to the limited analysis of human studies, articles published within the last 10 years, especially individuals aged 19 years and older. We used only free full text articles, restricted our studies to English-language articles on blood pressure control in hypertensive patients with salt restriction, and excluded articles related to other therapies from the review. Only eight of the studies were relevant to the study and were further assessed. Further research is needed to reach concrete conclusions.

## Conclusion

In conclusion, the profound impact of moderating sodium intake on proficiently managing blood pressure within the hypertensive population is unmistakable. This collective analysis unearths the potential dividends of implementing strategies that curtail salt consumption. Notably, a direct correlation emerges between adjusting dietary sodium and achieving proportional blood pressure reduction, spotlighting the indispensability of personalized interventions. Moreover, broader cardiovascular benefits become apparent with the adoption of a low-sodium diet, revealing intricate physiological responses that contribute to meticulous blood pressure regulation. Customized approaches underscore the promise of leveraging individual attributes and genetic influences to optimize outcomes. Mechanistic insights lay a robust foundation for endorsing the precision of reducing salt intake as an effective maneuver for blood pressure control. The horizons extend beyond blood pressure management, as evidenced by the crucial role of salt reduction in embracing the prevention of cardiovascular disease. Over time, longitudinal observations underscore the trajectory of blood pressure responses to salt reduction and tailored dietary plans, suggesting sustained benefits. In the real-world clinical setting, the pragmatic efficacy of interventions targeting sodium intake becomes evident, emphasizing tangible advantages. This confluence of evidence underscores the pivotal role of scaling back sodium consumption in achieving adept blood pressure management among those grappling with hypertension. Such an avenue holds exceptional promise for bolstering cardiovascular health through a network of interwoven mechanisms.

## Data Availability

All data produced in the present work are contained in the manuscript

